# Gaps in Vascular Evaluation Before Major Lower-Extremity Amputation Among Medicare Beneficiaries With Chronic Limb-Threatening Ischemia

**DOI:** 10.64898/2026.04.27.26351897

**Authors:** Samir K. Shah, Dan Neal, Khanjan B Shah, Terrie Vasilopoulos, Mark Segal, Salvatore T. Scali, Scott Berceli, Joel S. Weissman

## Abstract

**Background:** Guidelines recommend vascular specialist evaluation and revascularization consideration before major amputation in chronic limb-threatening ischemia (CLTI). Whether patients consistently receive pre-amputation vascular workup is poorly characterized nationally.

**Methods:** We conducted a retrospective cohort study of Medicare fee-for-service beneficiaries ≥66 years with CLTI undergoing incident major lower-extremity amputation (2021–2022) with ≥12 months continuous enrollment. Using claims in the 180 days preceding hospitalization for amputation, we classified patients into mutually exclusive pathway phenotypes: (A) no specialist, no imaging, no revascularization attempt; (B) specialist only, no revascularization attempt; (C) imaging, no revascularization attempt; or (D) revascularization attempted. Mixed-effects multinomial regression with hospital random intercepts identified predictors of phenotype membership. Post-amputation outcomes were compared across phenotypes.

**Results:** Among 10,666 patients (mean age 76.6 years; 35% female; 70% White, 21% Black), phenotype distribution was: A, 9.4%; B, 7.1%; C, 50.7%; D, 32.7%. Thus, 16.6% had no vascular imaging before amputation. Dementia (OR 2.0; 95% CI, 1.61–2.52), paralysis (OR 4.1; 2.62–6.34), and dual eligibility (OR 1.2; 1.01–1.42) were independently associated with phenotype A. Higher comorbidity burden was inversely associated with A (OR 0.49 for >6 vs 0–3 Elixhauser comorbidities). Phenotype A patients had lower 1-year mortality (40% vs 51% for D), fewer readmissions (90-day OR 0.54; 0.47–0.64), and lower costs (adjusted 50% lower at 180 days). Results were robust to acuity adjustment, exclusion of early deaths, and propensity-score matching (n=824 pairs). Phenotype A prevalence varied widely across hospital referral regions, ranging from 3% (Boston, Atlanta) to 16% (Little Rock) among regions with >100 patients.

**Conclusions:** One in six CLTI amputees had no vascular imaging before amputation. Patients without evaluation were characterized by cognitive impairment, functional limitation, lower healthcare engagement, and socioeconomic disadvantage rather than extreme medical complexity. Hospital-level variation suggests system-level interventions could address these gaps.

**WHAT IS KNOWN:**

- Prior studies have shown that 50–63% of Medicare patients with chronic limb-threatening ischemia undergo major amputation without receiving revascularization, with substantial racial and geographic disparities in pre-amputation vascular care.

**WHAT THE STUDY ADDS:**

- This study documents the extent to which CLTI patients proceed with amputation without first being evaluated by a vascular specialist, which suggests lack of guideline-recommended care.
- About 1 in 10 Medicare CLTI amputees had no vascular specialist contact and no vascular imaging in the 6 months before amputation.
- Patients reaching amputation without evaluation were characterized not by extreme medical complexity but by dementia, paralysis, depression, and dual eligibility—suggesting populations unable to self-advocate within the healthcare system.
- Substantial hospital-level and geographic variation (3–16% phenotype A prevalence across large hospital referral regions) indicates that system-level factors, not just patient characteristics, drive these gaps.

## INTRODUCTION

Major lower-extremity amputation for chronic limb-threatening ischemia (CLTI) carries perioperative mortality approaching 14% at 30 days and exceeding 48% at 1 year^1^. Clinical practice guidelines – including the 2019 Global Vascular Guidelines and 2024 ACC/AHA Peripheral Artery Disease Guidelines – require vascular specialist referral, hemodynamic testing, and anatomic imaging before major amputation can be considered appropriate^2,3^.

Despite these standards, prior studies have documented persistent underuse of pre-amputation vascular care. Goodney et al found that 54% of Medicare patients with CLTI had no revascularization procedures (e.g., bypass and stenting) in the year before amputation, with 6-fold regional variation^4,5^. Secemsky et al reported that 63% of CLTI amputees received low-intensity vascular care, associated with 21% higher 2-year mortality^6^. Other have further documented that 45% never saw a vascular specialist and 61% received no revascularization attempt^7^.

These studies reported rates of individual care components but did not classify patients into mutually exclusive pathway phenotypes mapping to the sequential logic of clinical care—referral, imaging, then treatment. Such a framework would identify the specific stage at which the evaluation process failed for each patient, enable direct comparison of outcomes across pathways, and provide insight into interventions to improve CLTI care. In this study, we used longitudinal Medicare fee-for-service claims to construct pathway phenotypes classifying each patient with CLTI undergoing major lower limb amputation by completeness of pre-amputation vascular evaluation, quantified patient and system factors associated with each phenotype, and compared post-amputation outcomes across pathways.

## METHODS

### Data Sources and Study Population

This retrospective cohort study used 2020–2022 Medicare fee-for-service claims from the Chronic Conditions Warehouse, including Inpatient, Outpatient, SNF, Carrier, and DME files; MedPAR; MBSF Base and Chronic Conditions files; and the Minimum Data Set. The files were based on a custom cohort designed to capture 100% of CLTI patients within the study time period. No formal sample size calculation was performed; all eligible Medicare fee-for-service beneficiaries meeting inclusion criteria during 2021–2022 were included. The study was approved by the University of Florida IRB.

We identified beneficiaries with major lower extremity amputation during 2021–2022 using International Classification of Disease, 10^th^ revision, Procedure Coding System (ICD-10-PCS) codes (**Supplemental Table S1**). Inclusion criteria were age ≥66 years, continuous FFS Parts A/B enrollment for ≥12 months before the index amputation, and CLTI diagnosis codes (**Supplemental Table S1**) on the index admission or within 12 months prior. We excluded patients with major amputation in the preceding 12 months.

### Pathway Phenotype Classification

Within the 180-day lookback window, we identified three hierarchical exposures from Carrier and Outpatient claims: (1) vascular subspecialty contact (CMS specialty codes 77, 94, C3 for vascular surgery, interventional radiology, and interventional cardiology); (2) vascular imaging (CPT 93922–93926, 75635, 73725, 75710–75716); and (3) endovascular or open revascularization attempts (**Supplemental Table S1** for ICD-10 codes). Each patient was classified into one mutually exclusive phenotype based on the 180-day lookback: **A** (no specialist, no imaging, no revascularization attempt), **B** (specialist only, no revascularization attempt), **C** (imaging, no revascularization attempt), or **D** (revascularization attempted). Patients with revascularization but no specialist/imaging claims were classified as D. Phenotype D was the only category consistent with clinical care guidelines for patients with potentially salvageable lower extremities. Phenotype assignment was repeated using a 90-day lookback as sensitivity analysis.

### Covariates and Contextual Markers

Covariates included age, sex, race/ethnicity, dual Medicare/Medicaid eligibility, Elixhauser mortality index^8^, and frailty. We used the claims-based frailty index (cFI), which is derived from 93 variables based on ICD diagnosis, CPT-4, and HCPCS codes and has been validated against outcomes including mortality, mobility impairment, and disability as well as clinical frailty measures^9^. We dichotomized frailty using the cutoff of ≥ 0.250 to represent mild versus moderate-to-severe frailty. In cases of severe limb-threatening disease or an unsalvageable limb, specialist evaluation and imaging may have been clinically inappropriate. To capture such cases, we used infection/sepsis (ICD-10-CM A40–A41, R65.2x), acute kidney injury (N17.*), and ICU use (revenue center codes 0200–0209) as markers of high index-admission acuity. Hospital attribution used the MedPAR provider identifier; hospital referral regions were identified for geographic analysis.

### Outcomes

Outcomes included all-cause mortality at 30, 90, and 365 days; readmission within 30 and 90 days; SNF utilization within 90 days; and total Medicare costs at 90 and 180 days. Costs were summed across inpatient, outpatient, SNF, carrier, and DME claims after the index admission, conditional on survival to the window endpoint.

### Statistical Analysis

Standard data quality procedures were applied prior to analysis, including removal of duplicate claim lines, exclusion of claims with service dates preceding beneficiary enrollment or following disenrollment, and verification of continuous enrollment periods using MBSF enrollment flags. All beneficiaries retained in the analytic cohort had complete records across linked CMS files. Phenotype predictors were modeled using mixed-effects multinomial logistic regression with hospital random intercepts (reference: phenotype D). Post-amputation outcomes used mixed-effects logistic regression (binary outcomes) and generalized linear models. All models included the full covariate set. Sensitivity analyses included: (1) additional adjustment for index-admission infection, AKI, and ICU use; (2) exclusion of deaths within 14 days; and (3) propensity-score matching of A to D on all covariates (caliper: absolute standardized mean difference <0.10 for all covariates). Kaplan-Meier curves were compared by log-rank test. Analyses were performed using the R software package (The R Foundation for Statistical Computing, v 4.5.1). Two-sided P<.05 was considered significant.

## RESULTS

### Cohort and Phenotype Distribution

The cohort comprised 10,666 patients (**Supplemental Figure S1**) with a mean age of 76.6 years and overall was 64.8% male, 69.6% White, and 21.4% Black. 69.3% diabetes with complications; 57.6% frailty index ≥0.25) In the 180-day lookback, 78% had vascular specialist contact, 82% had vascular imaging, and 33% underwent revascularization. Phenotype distribution was: A, 9.4% (n=1,006); B, 7.1% (n=761); C, 50.7% (n=5,406); D, 32.7% (n=3,493) (Table 1). With a 90-day lookback, phenotype A increased to 12.4% and D decreased to 27.6% (**Supplemental Table S2**).

**Table 1.**
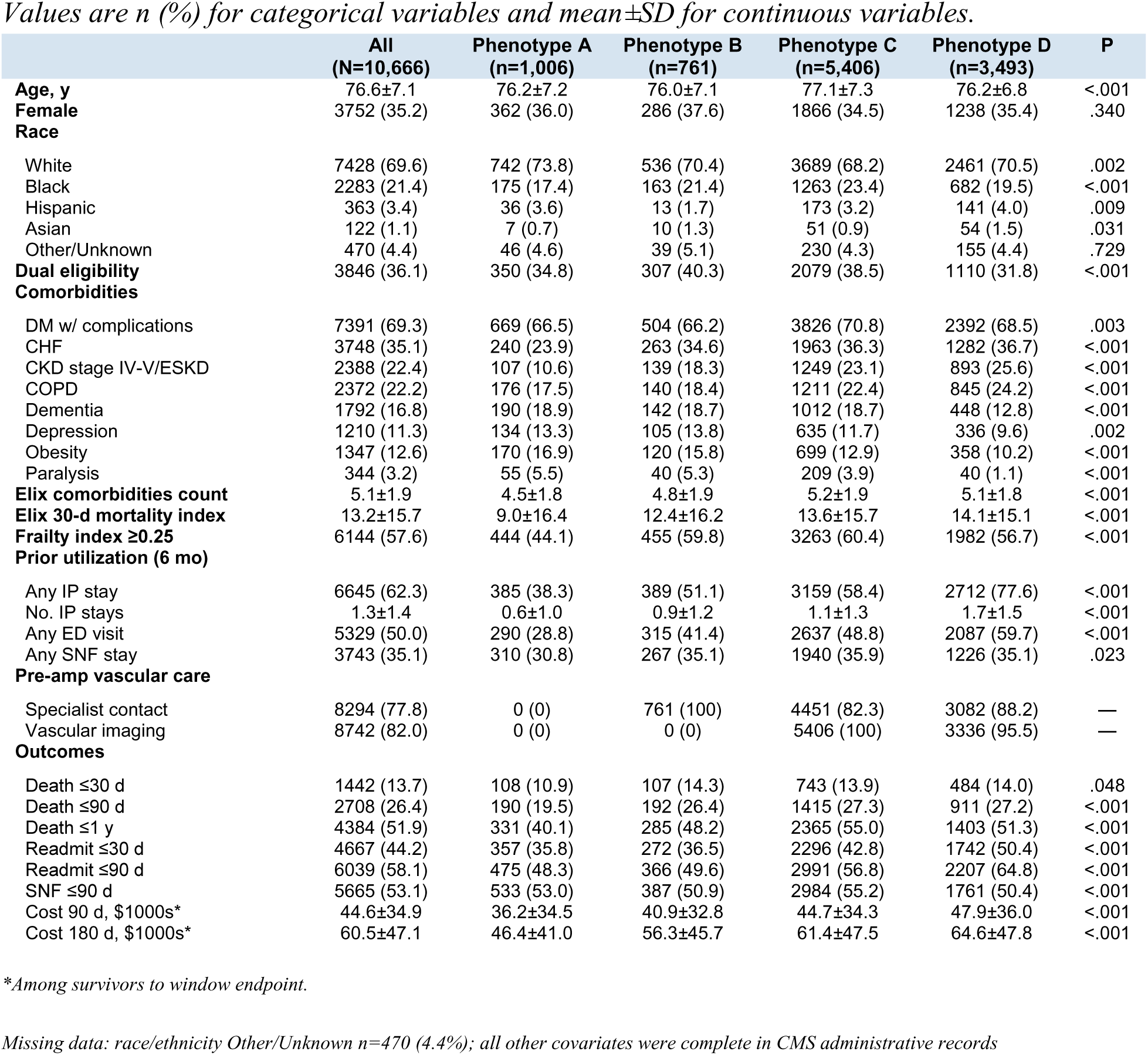
Cohort characteristics overall and by pathway phenotype. Values are n (%) or mean±SD. P from chi-square or ANOVA.

|  | All<br>(N=10,666) | Phenotype A<br>(n=1,006) | Phenotype B<br>(n=761) | Phenotype C<br>(n=5,406) | Phenotype D<br>(n=3,493) | P |
| --- | --- | --- | --- | --- | --- | --- |
| **Age, y** | 76.6±7.1 | 76.2±7.2 | 76.0±7.1 | 77.1±7.3 | 76.2±6.8 | <.001 |
| **Female** | 3752 (35.2) | 362 (36.0) | 286 (37.6) | 1866 (34.5) | 1238 (35.4) | .340 |
| **Race** |  |  |  |  |  |  |
| White | 7428 (69.6) | 742 (73.8) | 536 (70.4) | 3689 (68.2) | 2461 (70.5) | .002 |
| Black | 2283 (21.4) | 175 (17.4) | 163 (21.4) | 1263 (23.4) | 682 (19.5) | <.001 |
| Hispanic | 363 (3.4) | 36 (3.6) | 13 (1.7) | 173 (3.2) | 141 (4.0) | .009 |
| Asian | 122 (1.1) | 7 (0.7) | 10 (1.3) | 51 (0.9) | 54 (1.5) | .031 |
| Other/Unknown | 470 (4.4) | 46 (4.6) | 39 (5.1) | 230 (4.3) | 155 (4.4) | .729 |
| **Dual eligibility** | 3846 (36.1) | 350 (34.8) | 307 (40.3) | 2079 (38.5) | 1110 (31.8) | <.001 |
| **Comorbidities** |  |  |  |  |  |  |
| DM w/ complications | 7391 (69.3) | 669 (66.5) | 504 (66.2) | 3826 (70.8) | 2392 (68.5) | .003 |
| CHF | 3748 (35.1) | 240 (23.9) | 263 (34.6) | 1963 (36.3) | 1282 (36.7) | <.001 |
| CKD stage IV-V/ESKD | 2388 (22.4) | 107 (10.6) | 139 (18.3) | 1249 (23.1) | 893 (25.6) | <.001 |
| COPD | 2372 (22.2) | 176 (17.5) | 140 (18.4) | 1211 (22.4) | 845 (24.2) | <.001 |
| Dementia | 1792 (16.8) | 190 (18.9) | 142 (18.7) | 1012 (18.7) | 448 (12.8) | <.001 |
| Depression | 1210 (11.3) | 134 (13.3) | 105 (13.8) | 635 (11.7) | 336 (9.6) | .002 |
| Obesity | 1347 (12.6) | 170 (16.9) | 120 (15.8) | 699 (12.9) | 358 (10.2) | <.001 |
| Paralysis | 344 (3.2) | 55 (5.5) | 40 (5.3) | 209 (3.9) | 40 (1.1) | <.001 |
| **Elix comorbidities count** | 5.1±1.9 | 4.5±1.8 | 4.8±1.9 | 5.2±1.9 | 5.1±1.8 | <.001 |
| **Elix 30-d mortality index** | 13.2±15.7 | 9.0±16.4 | 12.4±16.2 | 13.6±15.7 | 14.1±15.1 | <.001 |
| **Frailty index ≥0.25** | 6144 (57.6) | 444 (44.1) | 455 (59.8) | 3263 (60.4) | 1982 (56.7) | <.001 |
| **Prior utilization (6 mo)** |  |  |  |  |  |  |
| Any IP stay | 6645 (62.3) | 385 (38.3) | 389 (51.1) | 3159 (58.4) | 2712 (77.6) | <.001 |
| No. IP stays | 1.3±1.4 | 0.6±1.0 | 0.9±1.2 | 1.1±1.3 | 1.7±1.5 | <.001 |
| Any ED visit | 5329 (50.0) | 290 (28.8) | 315 (41.4) | 2637 (48.8) | 2087 (59.7) | <.001 |
| Any SNF stay | 3743 (35.1) | 310 (30.8) | 267 (35.1) | 1940 (35.9) | 1226 (35.1) | .023 |
| **Pre-amp vascular care** |  |  |  |  |  |  |
| Specialist contact | 8294 (77.8) | 0 (0) | 761 (100) | 4451 (82.3) | 3082 (88.2) | — |
| Vascular imaging | 8742 (82.0) | 0 (0) | 0 (0) | 5406 (100) | 3336 (95.5) | — |
| **Outcomes** |  |  |  |  |  |  |
| Death ≤30 d | 1442 (13.7) | 108 (10.9) | 107 (14.3) | 743 (13.9) | 484 (14.0) | .048 |
| Death ≤90 d | 2708 (26.4) | 190 (19.5) | 192 (26.4) | 1415 (27.3) | 911 (27.2) | <.001 |
| Death ≤1 y | 4384 (51.9) | 331 (40.1) | 285 (48.2) | 2365 (55.0) | 1403 (51.3) | <.001 |
| Readmit ≤30 d | 4667 (44.2) | 357 (35.8) | 272 (36.5) | 2296 (42.8) | 1742 (50.4) | <.001 |
| Readmit ≤90 d | 6039 (58.1) | 475 (48.3) | 366 (49.6) | 2991 (56.8) | 2207 (64.8) | <.001 |
| SNF ≤90 d | 5665 (53.1) | 533 (53.0) | 387 (50.9) | 2984 (55.2) | 1761 (50.4) | <.001 |
| Cost 90 d, \$1000s* | 44.6±34.9 | 36.2±34.5 | 40.9±32.8 | 44.7±34.3 | 47.9±36.0 | <.001 |
| Cost 180 d, \$1000s* | 60.5±47.1 | 46.4±41.0 | 56.3±45.7 | 61.4±47.5 | 64.6±47.8 | <.001 |

\*Among survivors to window endpoint.
Missing data: race/ethnicity Other/Unknown n=470 (4.4%); all other covariates were complete in CMS administrative records

### Characteristics by Phenotype

Phenotype A patients were the least medically complex (**Table 1**). Compared with patients in Phenotype D, they had a lower Elixhauser mortality index (9.0 vs 14.1), lower frailty (44.1% vs 56.7%), and lower rates of congestive heart failure (23.9% vs 36.7%), and chronic kidney disease stages IV-V or end-stage kidney disease (10.6% vs 25.6%) (all p<.001). Conversely, phenotype A had high rates of dementia (18.9% vs 12.8%-18.7% for other phenotypes), depression (13.3% vs 9.6%-13.8%), obesity (16.9% vs 10.2%-15.8%), and paralysis (5.5% vs 1.1%-5.3%) (all p≤.002). Prior healthcare utilization was dramatically lower: only 38.3% had any inpatient stay in the prior 6 months (vs 77.6% for D; mean 0.6 vs 1.7 prior hospitalizations). ED visits showed a similar pattern (28.8% vs 59.7%).

Seventy-seven percent of patients had infection, AKI, and/or ICU use on the index admission. Of these features, phenotype A patients were more likely to present with infection alone (34.9% vs 23.1% for D) but less likely to have ICU stays (18.4% vs 37.0%) (**Supplemental Table S3**).

### Predictors of Phenotype Membership

The strongest independent predictors of phenotype A (vs D) were paralysis (OR 4.1; 95% CI, 2.6–6.3), severe liver disease (OR 4.5; 1.5–13.9), dementia (OR 2.0; 1.6–2.5), psychoses (OR 1.9; 1.2–3.1), drug abuse (OR 1.9; 1.1–3.3), obesity (OR 1.6; 1.3–1.9), depression (OR 1.5; 1.2–1.8), and dual eligibility (OR 1.4; 1.2–1.6) (**Table 2**, **Figure 2**). Increasing Elixhauser burden showed a dose-response inverse association: OR 0.77, 0.63, 0.62, and 0.49 for 4, 5, 6, and >6 comorbidities versus 0–3 (all P<.001). Frailty was associated with lower odds of phenotype A (OR 0.65; 0.6–0.8) but higher odds of C (OR 1.1; 1.0–1.2). Black race was associated with higher odds of C (OR 1.2; 1.04–1.29) but not A. Dual eligibility was associated with all non-D phenotypes.

**Figure 1.**
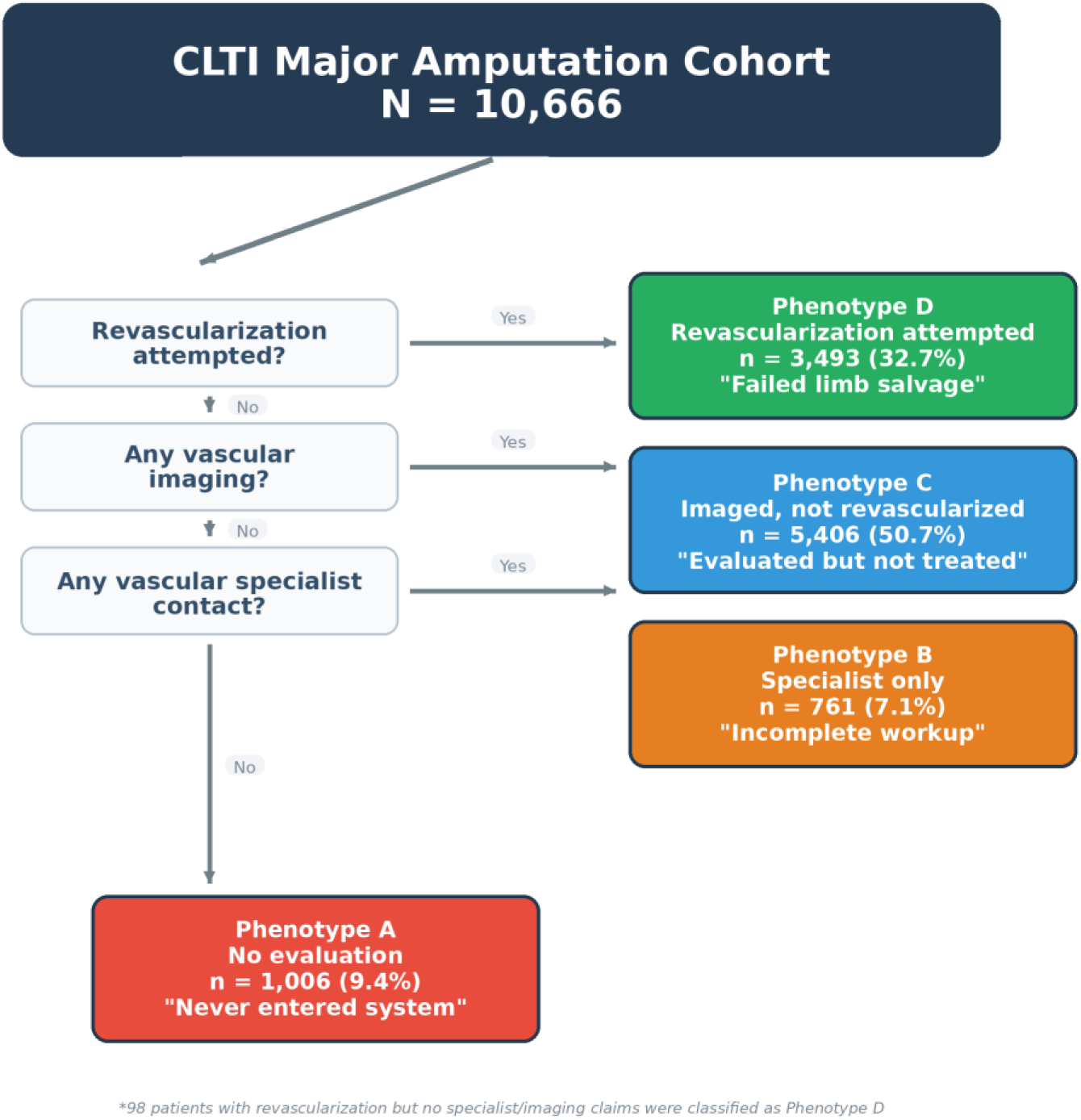
Pathway phenotype schematic. Four mutually exclusive phenotypes (A–D) defined by completeness of vascular evaluation in the 180-day lookback window, with operational definitions and sample sizes.

**Figure 2.**
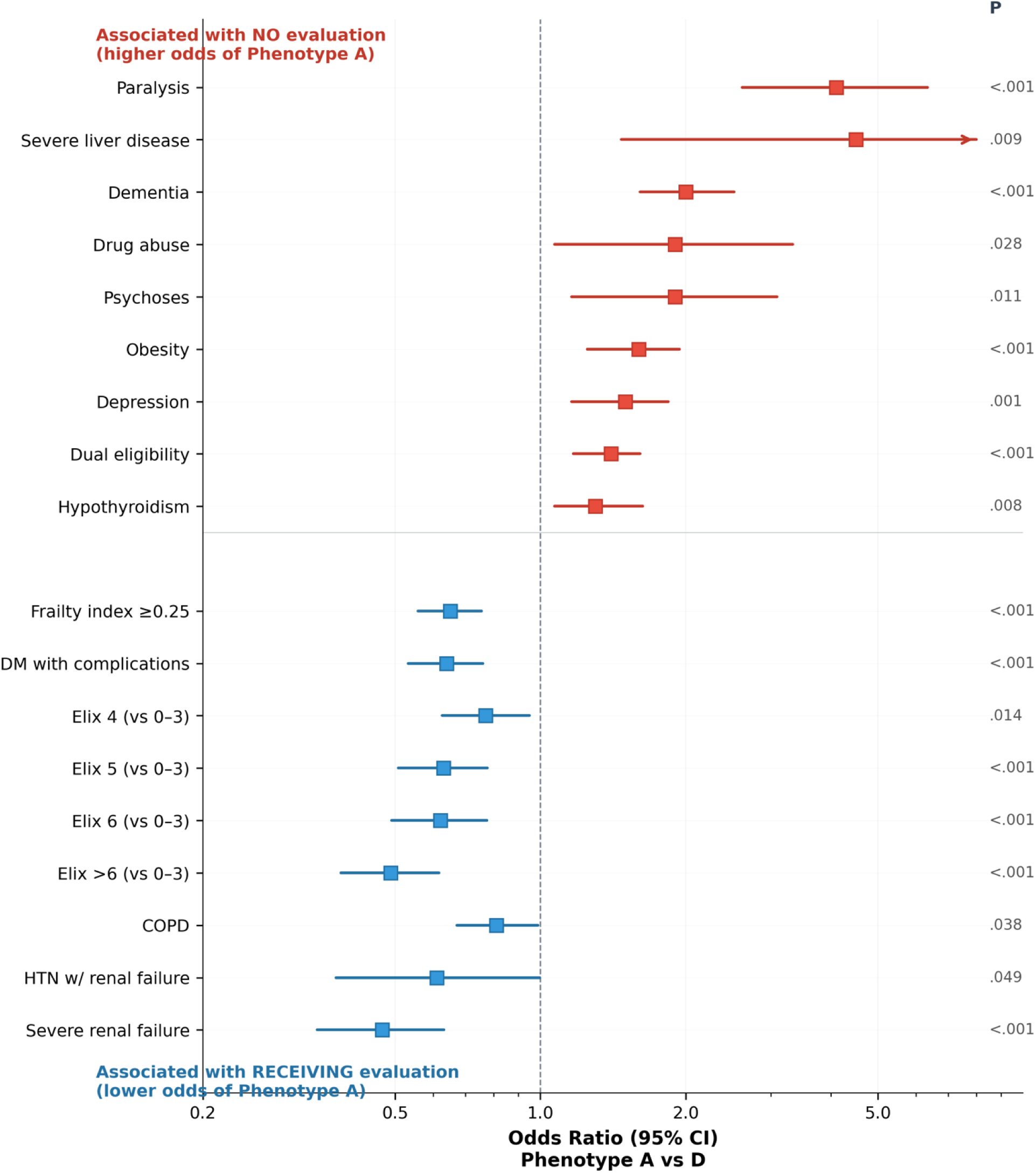
Factors associated with no vascular evaluation (Phenotype A). Forest plot of odds ratios from mixed-effects multinomial logistic regression (reference: Phenotype D) with hospital random intercept. Red markers indicate factors associated with higher odds of reaching amputation without evaluation; blue markers indicate factors associated with having received evaluation. Error bars represent 95% confidence intervals on a logarithmic scale.

**Table 2.**
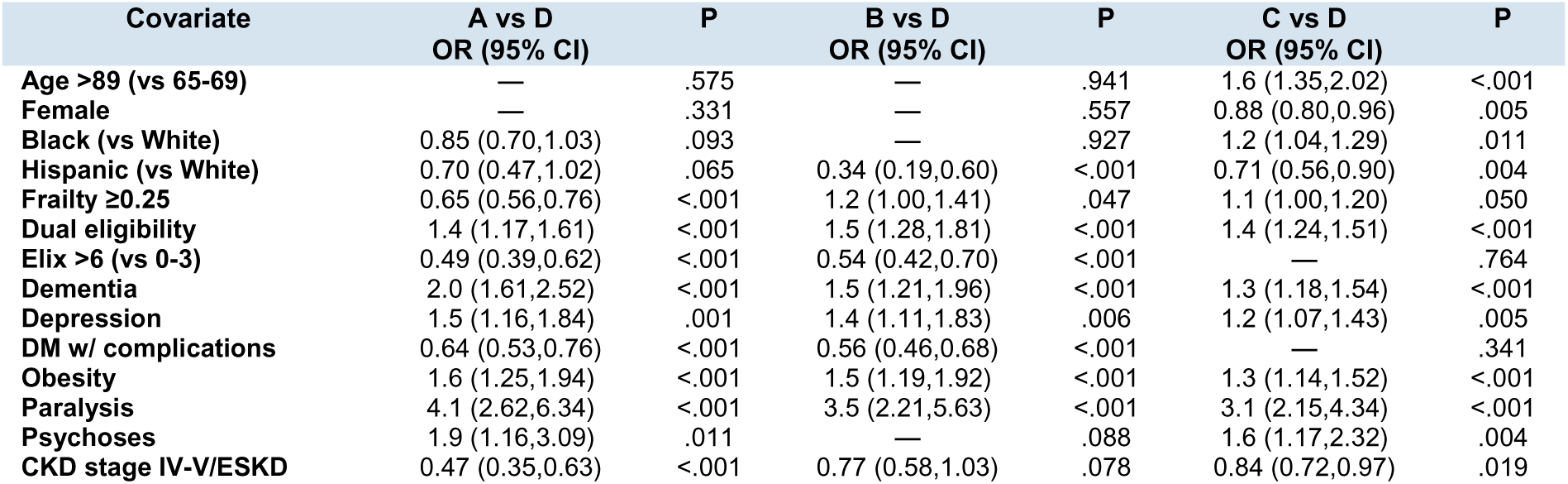
Mixed-effects multinomial regression: predictors of pathway phenotype. Reference: D. Hospital random intercept included. ORs (95% CI) shown for comparisons with P<.10.

### Hospital and Geographic Variation

There was moderate between-hospital variation in risk-adjusted phenotype A probability across 2,062 hospitals. Among 20 HRRs with >100 patients, phenotype A prevalence ranged from 3% (Atlanta, Boston, St. Louis) to 16% (Little Rock) and 15% (Washington, DC) (**Supplemental Table S4**).

### Post-Amputation Outcomes

One-year mortality was 40.1% for A, 48.2% for B, 55.0% for C, and 51.3% for D (Figure 3; log-rank P<.0001). Kaplan-Meier curves showed sustained separation favoring phenotype A. Readmission rates were substantially lower for A: 30-day 35.8% vs 50.4% for D; 90-day 48.3% vs 64.8% (P<.0001). Among 180-day survivors, mean costs were $46,400 for A versus $64,600 for D (**Table 3**).

**Figure 3.**
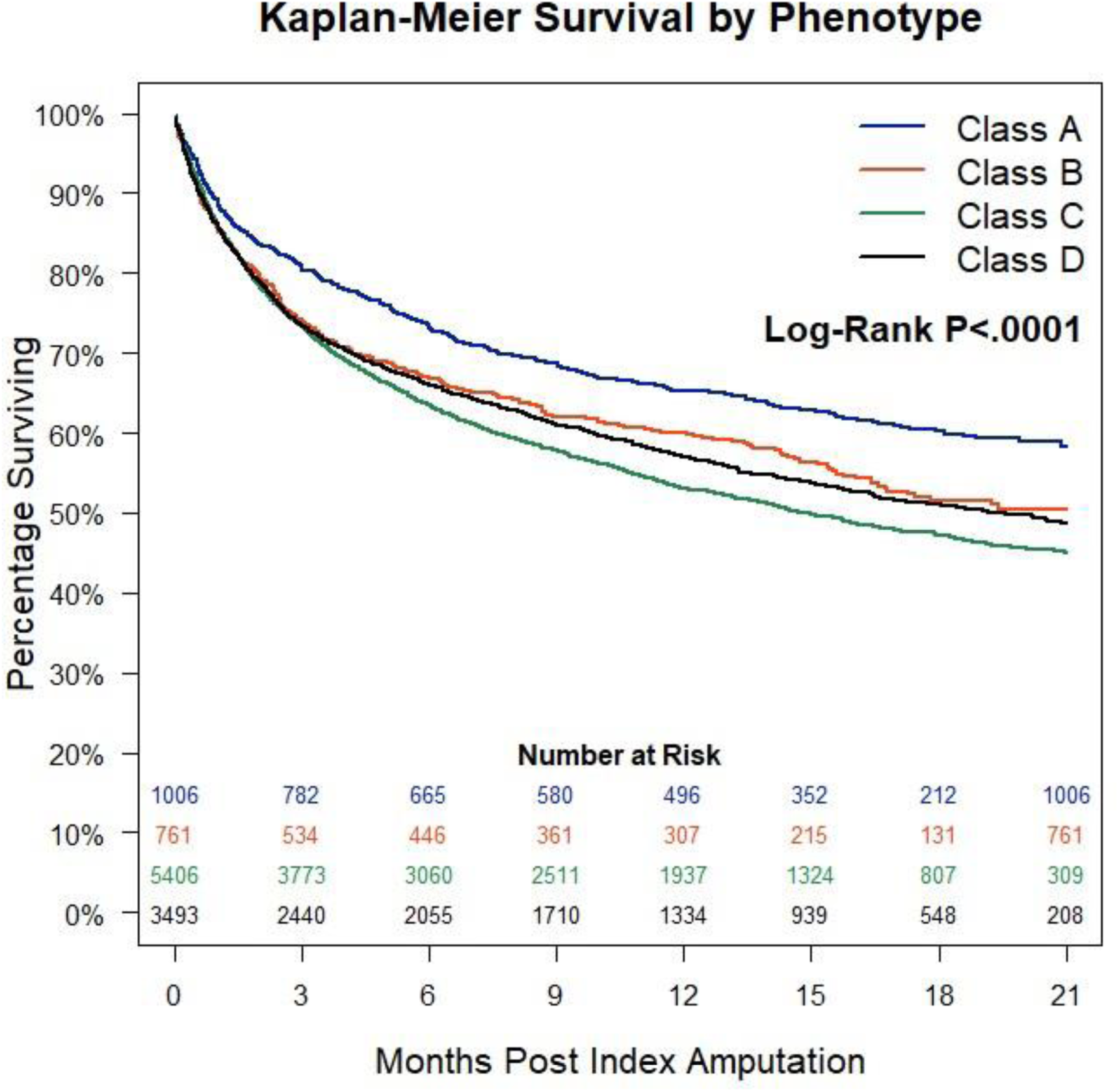
Kaplan-Meier survival by pathway phenotype. All-cause survival from index amputation through 21 months, stratified by phenotype (log-rank P<.0001). Number at risk shown.

**Table 3.** Post-amputation outcomes by phenotype: adjusted models. Reference: D. Original model and model adjusted for infection, AKI, ICU.

| Outcome | Original Effect (95% CI) | P | Adjusted for Inf/AKI/ICU | P |
| --- | --- | --- | --- | --- |
| **30-day death** |  |  |  |  |
| A vs D | — | .444 | — | .693 |
| B vs D | — | .569 | — | .359 |
| C vs D | — | .979 | — | .425 |
| **90-day death** |  |  |  |  |
| A vs D | 0.80 (0.66,0.97) | .025 | 0.82 (0.68,1.00) | .053 |
| B vs D | — | .993 | — | .759 |
| C vs D | — | .833 | — | .593 |
| **1-year death** |  |  |  |  |
| A vs D | 0.82 (0.68,0.98) | .030 | 0.84 (0.70,1.00) | .053 |
| B vs D | — | .299 | — | .363 |
| C vs D | 1.1 (1.02,1.27) | .019 | 1.2 (1.05,1.30) | .001 |
| **30-day readmission** |  |  |  |  |
| A vs D | 0.56 (0.48,0.66) | <.001 | 0.57 (0.48,0.67) | <.001 |
| B vs D | 0.62 (0.52,0.74) | <.001 | 0.63 (0.53,0.75) | <.001 |
| C vs D | 0.79 (0.72,0.86) | <.001 | 0.79 (0.72,0.87) | <.001 |
| **90-day readmission** |  |  |  |  |
| A vs D | 0.55 (0.47,0.64) | <.001 | 0.54 (0.47,0.64) | <.001 |
| B vs D | 0.59 (0.50,0.70) | <.001 | 0.59 (0.50,0.70) | <.001 |
| C vs D | 0.76 (0.69,0.84) | <.001 | 0.76 (0.69,0.83) | <.001 |
| **90-day costs (survivors)** |  |  |  |  |
| A vs D | 48% lower | <.001 | 48% lower | <.001 |
| B vs D | 26% lower | <.001 | 26% lower | <.001 |
| C vs D | — | .422 | — | .422 |
| **180-day costs (survivors)** |  |  |  |  |
| A vs D | 50% lower | <.001 | 50% lower | <.001 |
| B vs D | 24% lower | .001 | 24% lower | .001 |
| C vs D | — | .337 | — | .337 |

**Figure 4.**
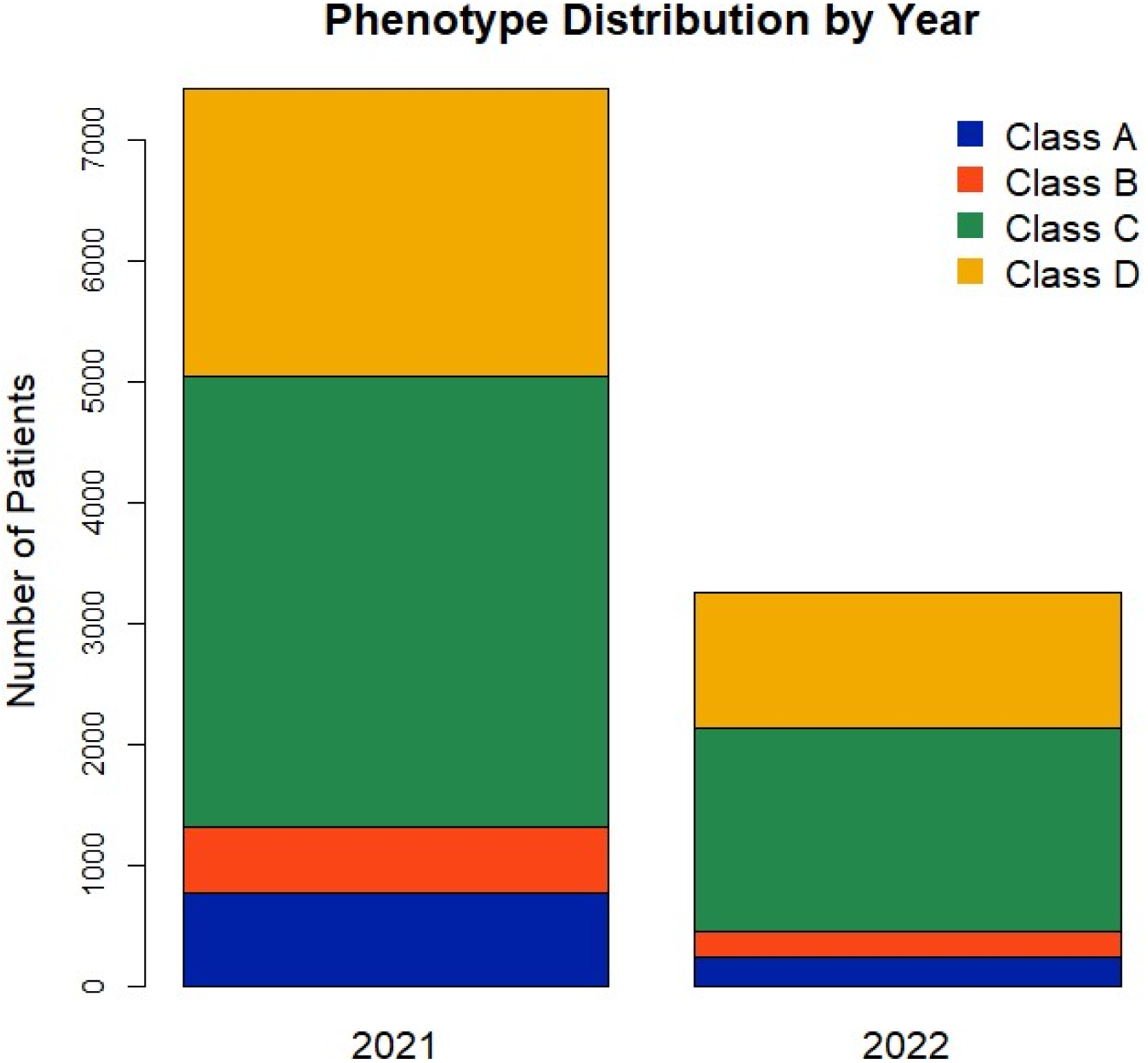
Phenotype distribution by year of index amputation (2021 vs 2022).

In adjusted models after excluding deaths within 14 days of amputation (**Table 3**), 90-day mortality was OR 0.8 (0.7–1.0; P=.056) and 1-year mortality OR 0.8 (0.7–1.0; P=.053) for A versus D after adjusting for index-admission acuity markers. Phenotype C had significantly higher 1-year mortality than D (OR 1.2; 1.1–1.3; P=.011). Readmission differences were highly robust: 30-day OR 0.6 (0.5–0.7; P<.0001) and 90-day OR 0.6 (0.5–0.6; P<.0001) for A versus D. Adjusted 180-day costs were 50% lower for A versus D (P<.0001).

Among patients with imaging (phenotypes C and D), time from first imaging to amputation was shorter for C (median 49 days; IQR, 10–110) than D (median 79 days; IQR, 27–134; P<.0001), with 25% of C patients imaged within 10 days of amputation.

### Sensitivity Analyses

Excluding deaths within 14 days (n=815; phenotype distribution unchanged) produced virtually identical results (**Supplemental Table S2**). Propensity-matched analysis (824 pairs; **Table 4**) confirmed: 1-year mortality 41.0% vs 46.5% (OR 0.8; P=.038), 30-day readmission 35.0% vs 50.6% (OR 0.5; P<.0001), and 180-day costs 72% lower for A, p<.0001. The E-value for the 30-day readmission OR was 2.1 (95% CI, 1.8–2.4). Notably, 20.6% of phenotype A patients had no infection, AKI, or ICU use on the index admission.

**Table 4.** Propensity-score matched comparison, phenotype A vs D (n=824 pairs).

| Outcome | Class A (n=824) | Class D (n=824) | Effect (P value) |
| --- | --- | --- | --- |
| 30-day death | 92 (11.3) | 85 (10.4) | P = .577 |
| 90-day death | 159 (19.9) | 185 (23.3) | OR 0.82 (0.65,1.03); P=.082 |
| 1-year death | 279 (41.0) | 297 (46.5) | OR 0.80 (0.65,0.99); P=.038 |
| 30-day readmission | 286 (35.0) | 413 (50.6) | OR 0.52 (0.43,0.64); P<.0001 |
| 90-day readmission | 382 (47.5) | 529 (65.7) | OR 0.47 (0.39,0.58); P<.0001 |
| SNF stay ≤90 d | 466 (56.6) | 433 (52.5) | OR 1.2 (0.98,1.42); P=.088 |
| 90-d costs, \$1000s | 3.3±4.3 | 4.9±5.7 | 72% lower; P<.0001 |
| 180-d costs, \$1000s | 4.8±7.0 | 7.4±15.5 | 72% lower; P<.0001 |

## DISCUSSION

In this national cohort of 10,666 Medicare CLTI amputees, we found that 16.6% had no vascular imaging and 9.4% had neither specialist contact nor imaging in the 180 days before amputation. We identified that unevaluated patients were characterized not by extreme medical complexity but by cognitive impairment, functional limitation, lower healthcare engagement, and socioeconomic disadvantage. Substantial hospital-level and geographic variation suggests these gaps may be driven in part by system-level factors amenable to intervention.

Our phenotype approach extends prior work by Goodney et al, Secemsky et al, and Ramadan et al by identifying the specific stage at which the evaluation cascade failed^4–7^. Phenotype A (no specialist, no imaging, no revascularization attempt) represents a referral or access gap (no system entry); phenotype B (specialist only, no revascularization attempt) suggests incomplete workup; phenotype C (imaging, no revascularization attempt) captures evaluation without treatment (potentially appropriate for no-option anatomy or patient preference); and phenotype D (revascularization attempted) represents failed limb salvage.

We speculate that the lower mortality among those in phenotype A reflects better health and lower engagement with healthcare, which could reflect underlying socioeconomic disadvantages or poor health literacy. These patients had fewer comorbidities (Elixhauser 4.5 vs 5.1), lower frailty, and dramatically less prior healthcare utilization (0.6 vs 1.7 prior hospitalizations). The dose-response relationship between comorbidity burden and phenotype A (OR 0.49 for >6 vs 0–3 comorbidities) confirms that lower medical complexity characterizes this group. We interpret this as a phenotype of relatively healthy but healthcare-disengaged/disadvantaged patients.

Higher rates of dual eligibility (34.8% vs 31.8%) support this interpretation. Lower readmission rates and costs further reflect continued lower healthcare engagement after amputation. These findings should not be interpreted as evidence that absence of vascular evaluation improves outcomes; rather, they reflect a population whose lower baseline medical complexity and healthcare engagement accounts for their relatively better post-amputation survival.

The strongest predictors of absent evaluation—dementia (OR 2.0), paralysis (OR 4.1), psychoses (OR 1.9), depression (OR 1.5)—are conditions impairing patients’ ability to self-advocate. This aligns with evidence that frail CLTI patients are 22% less likely to receive revascularization^10^. Our data suggest these patients are not just triaged to amputation after evaluation but that they reach amputation without evaluation entirely.

The 9.4% in phenotype A represent a potentially preventable gap: patients who reached irreversible limb loss without the minimum evaluation to determine if salvage was possible. Proactive referral systems—automated EHR flags for CLTI patients not seen by a vascular specialist—could reduce reliance on patient self-advocacy. The substantial hospital-level variation supports institutional quality measures for pre-amputation vascular evaluation.

### Limitations

Claims cannot capture clinical rationale for decisions, informal evaluations, or care outside Medicare FFS (VA, Medicare Advantage). The study period overlaps the COVID-19 pandemic, which may have affected care patterns. As with all studies using administrative claims, diagnoses and procedures are recorded for billing purposes rather than research, introducing potential misclassification in both CLTI identification and phenotype assignment. Additionally, changes in ICD-10 coding practices over the 2021–2022 study period may have affected eligibility classification over time, potentially biasing phenotype prevalence estimates. Incomplete capture of non-covered services (e.g., VA care) may also result in undermeasurement of actual care received. Home health claims were unavailable for cost analyses. Residual confounding from unmeasured functional status and patient preferences cannot be excluded; the E-value of 2.1 for readmission suggests moderate unmeasured confounding could account for some associations. Pathway phenotypes are claims-based proxies; phenotype C includes patients with no-option anatomy, prohibitive risk, or an informed choice against intervention.

## CONCLUSIONS

Among 10,666 Medicare CLTI amputees, 16.6% had no vascular imaging and 9.4% had no specialist contact or imaging before amputation. The unevaluated population was characterized by cognitive impairment, functional limitation, and socioeconomic disadvantage rather than extreme medical complexity, with substantial hospital-level variation. These findings support targeted system-level interventions to ensure vascular evaluation before irreversible limb loss, with particular attention to socioeconomically disadvantaged patients and other groups characterized by impaired self-advocacy.

## Data Sharing Statement

The data used in this study are available from the Centers for Medicare and Medicaid Services (CMS) Chronic Conditions Warehouse. Due to data use agreement restrictions, the analytic dataset cannot be shared directly. The analytic code is available from the corresponding author upon reasonable request.

## IRB Statement

This study was approved by the University of Florida Institutional Review Board.

**Supplemental Table S1.** Procedure and diagnosis code lists for major lower-extremity amputation, CLTI, vascular imaging, and revascularization.

**Supplemental Table S2.** Phenotype distribution with 90-day lookback.

**Supplemental Table S3.** Acuity markers by phenotype; outcome models excluding early deaths.

**Supplemental Table S4.** Phenotype A prevalence by HRR (>100 patients).

**Supplemental Figure S1.** Cohort flow diagram.

